## Supplemental Tables and Figures for "Association of computed tomography scan–assessed body composition with immune and PI3K/AKT pathway proteins in distinct breast cancer tumor components"

**Supplementary Materials**

Supplemental Table S1. Time interval (days) between computed tomography (CT) scan procedure and breast cancer diagnosis and between CT scan and surgery, the time when the tumor samples were obtained

|  | **No.** | **Median** | **Interquartile range** |
| --- | --- | --- | --- |
| **Time between CT and diagnosis** |  |  |  |
| CT performed before diagnosis | 20 | 173 | 74 – 344 |
| CT performed after diagnosis | 32 | 111 | 35 – 592 |
| **Time between CT and surgery^a^** |  |  |  |
| CT performed before surgery | 25 | 121 | 23 – 253 |
| CT performed after surgery | 24 | 173 | 29 – 346 |

^a^ Three patients had missing data on the date of surgery.

Supplemental Table S2. Tissue localization sites and Hounsfield unit cut points used for computed tomography scan annotations

| **Tissue** | **Localization sites** | **Hounsfield unit, lower limit** | **Hounsfield unit, upper limit** |
| --- | --- | --- | --- |
| Subcutaneous adipose tissue (SAT) | Subcutaneous fat, skin | -190 | -30 |
| Intermuscular Adipose tissue (IMAT) | Inter-muscular fat, marrow^a^ | -190 | -30 |
| Visceral Adipose tissue (VAT) | Fat around organs | -150 | -50 |
| Very low-density muscle (VLDM) | Muscle-fat boarders, skin, marrow | -29 | 0 |
| Low density muscle (LDM) | Inter- & intra-muscular, skin, muscle-fat, and marrow borders | 0 | 35 |
| Normal density muscle (NDM) | Muscle, skin, marrow | 35 | 101 |
| High density muscle (HDM) | Bone-muscle border, intra-muscular | 101 | 151 |
| Very high-density muscle (VHDM) | Bone-muscle and bone-marrow boarders | 151 | 200 |

^a^ Marrow was excluded from annotations.

Supplemental Table S4. Areas of specific body composition components, total area (cm^2^), and waist circumference (cm) by body composition phenotype

| Body composition phenotype | Number of Observations | Variable | Mean | SD | Minimum | Maximum |
| --- | --- | --- | --- | --- | --- | --- |
| Adequate | 22 | VAT | 98.49 | 59.67 | 8.86 | 216.80 |
|  |  | SAT | 251.92 | 119.05 | 2.95 | 440.10 |
|  |  | IMAT | 17.03 | 35.35 | 0.95 | 168.50 |
|  |  | VLDM | 17.46 | 10.76 | 6.94 | 56.28 |
|  |  | LDM | 40.60 | 17.30 | 19.20 | 99.56 |
|  |  | NDM | 65.50 | 25.78 | 0.80 | 103.70 |
|  |  | HDM | 2.95 | 1.42 | 0.55 | 5.24 |
|  |  | VHDM | 2.96 | 1.38 | 0.56 | 6.00 |
|  |  | TAT | 367.44 | 138.54 | 65.66 | 522.82 |
|  |  | TSM | 129.47 | 11.17 | 116.54 | 159.63 |
|  |  | Total area | 496.91 | 140.35 | 194.19 | 666.79 |
|  |  | WC | 97.09 | 13.48 | 71.09 | 119.70 |
| High adiposity | 13 | VAT | 207.97 | 76.28 | 77.75 | 358.00 |
|  |  | SAT | 447.15 | 102.60 | 321.50 | 670.40 |
|  |  | IMAT | 9.97 | 5.04 | 2.55 | 22.59 |
|  |  | VLDM | 23.05 | 9.64 | 12.50 | 49.84 |
|  |  | LDM | 50.22 | 10.44 | 28.15 | 69.81 |
|  |  | NDM | 66.03 | 24.84 | 26.86 | 117.00 |
|  |  | HDM | 4.92 | 2.26 | 1.73 | 8.44 |
|  |  | VHDM | 4.27 | 0.92 | 2.92 | 5.66 |
|  |  | TAT | 665.09 | 114.59 | 535.96 | 872.37 |
|  |  | TSM | 148.49 | 18.68 | 120.58 | 179.30 |
|  |  | Total area | 813.58 | 122.15 | 659.34 | 1040.67 |
|  |  | WC | 110.73 | 14.79 | 78.99 | 141.10 |
| Low muscle | 14 | VAT | 82.12 | 61.51 | 8.32 | 219.60 |
|  |  | SAT | 233.28 | 82.45 | 93.88 | 368.90 |
|  |  | IMAT | 7.04 | 4.59 | 0.80 | 15.87 |
|  |  | VLDM | 12.59 | 5.09 | 5.19 | 24.73 |
|  |  | LDM | 30.45 | 9.75 | 16.33 | 47.14 |
|  |  | NDM | 56.10 | 13.99 | 38.23 | 86.40 |
|  |  | HDM | 3.91 | 1.28 | 1.91 | 5.85 |
|  |  | VHDM | 3.26 | 1.12 | 1.12 | 6.07 |
|  |  | TAT | 322.44 | 131.76 | 104.21 | 522.15 |
|  |  | TSM | 106.30 | 7.19 | 96.70 | 115.46 |
|  |  | Total area | 428.74 | 134.25 | 206.24 | 636.36 |
|  |  | WC | 91.99 | 6.83 | 79.00 | 102.70 |
| High adiposity with Low muscle | 3 | VAT | 151.23 | 48.37 | 109.20 | 204.10 |
|  |  | SAT | 406.40 | 77.40 | 317.80 | 460.90 |
|  |  | IMAT | 15.41 | 11.62 | 2.00 | 22.38 |
|  |  | VLDM | 18.86 | 9.40 | 8.72 | 27.27 |
|  |  | LDM | 33.63 | 13.27 | 19.76 | 46.20 |
|  |  | NDM | 43.83 | 29.83 | 23.24 | 78.04 |
|  |  | HDM | 3.59 | 2.59 | 2.02 | 6.58 |
|  |  | VHDM | 3.33 | 0.67 | 2.68 | 4.03 |
|  |  | TAT | 573.05 | 29.25 | 544.28 | 602.76 |
|  |  | TSM | 103.23 | 11.51 | 90.42 | 112.70 |
|  |  | Total area | 676.28 | 22.41 | 650.85 | 693.18 |
|  |  | WC | 100.93 | 9.34 | 90.18 | 107.10 |

Abbreviations: VAT, visceral adipose tissue; SAT, subcutaneous adipose tissue; IMAT, intermuscular adipose tissue; VLDM, very low-density muscle; LDM, low-density muscle; NDM, normal-density muscle; HDM, high-density muscle; VHDM, very high-density muscle; TAT, total adipose tissue; TSM, total skeleton muscle tissue; WC, waist circumference.

Supplemental Table S5. Body composition group by patient and clinical characteristics

| Characteristic | Body composition type (N = 52) | | | |
| --- | --- | --- | --- | --- |
|  | Adequate | High adiposity | Low muscle | Low muscle/high adiposity |
| N | 22 | 13 | 14 | 3 |
| Age, mean (SD), years | 54.5 (10.9) | 59.1 (9.1) | 58.3 (11.4) | 57.7 (10.2) |
| Age category, % |  |  |  |  |
| <55 | 54.6 | 30.8 | 42.9 | 33.3 |
| ≥55 | 45.4 | 69.2 | 57.1 | 66.7 |
| Race, % |  |  |  |  |
| White | 77.3 | 84.6 | 85.7 | 100.0 |
| Black | 22.7 | 15.4 | 14.3 | 0.0 |
| AJCC stage, % |  |  |  |  |
| I | 31.8 | 30.8 | 35.7 | 33.3 |
| II | 50.0 | 61.5 | 42.9 | 33.3 |
| III | 18.2 | 7.7 | 21.4 | 33.3 |
| Tumor grade, % |  |  |  |  |
| I | 4.6 | 30.8 | 0.0 | 0.0 |
| II | 50.0 | 46.1 | 57.1 | 100.0 |
| III | 45.4 | 23.1 | 42.9 | 0.0 |
| Tumor size, % |  |  |  |  |
| <25 mm | 36.4 | 61.5 | 50.0 | 33.3 |
| ≥25 mm | 40.9 | 30.8 | 28.6 | 0.0 |
| Missing | 22.7 | 7.7 | 21.4 | 66.7 |
| ER status, % |  |  |  |  |
| Positive | 81.8 | 100.0 | 85.7 | 100.0 |
| Negative | 18.2 | 0.0 | 14.3 | 0.0 |
| PR status, % |  |  |  |  |
| Positive | 68.2 | 100.0 | 85.7 | 100.0 |
| Negative | 31.8 | 0.0 | 14.3 | 0.0 |
| HER2 status, % |  |  |  |  |
| Positive | 31.8 | 0.0 | 21.4 | 0.0 |
| Negative | 68.2 | 100.0 | 78.6 | 100.0 |

Abbreviations: AJCC, American Joint Committee on Cancer; ER; ER, estrogen receptor; PR, progesterone receptor

Supplemental Table S6. Percentages of obesity and sarcopenia in each body composition type

| Body composition type | Overall^a^ | Adequate | High adiposity | Low muscle | High adiposity with low muscle |
| --- | --- | --- | --- | --- | --- |
| Total | 43 | 18 | 11 | 11 | 3 |
| Obesity |  |  |  |  |  |
| No (BMI <30 kg/m^2^) | 23 (53.5%) | 10 (55.6%) | 2 (18.2%) | 9 (81.8%) | 2 (66.7%) |
| Yes (BMI ≥30 kg/m^2^) | 20 (46.5%) | 8 (44.4%) | 9 (81.8%) | 2 (18.2%) | 1 (33.3%) |
| Sarcopenia^b^ |  |  |  |  |  |
| No | 38 (88.4%) | 17 (94.4%) | 11 (100%) | 8 (72.7%) | 2 (66.7%) |
| Yes | 5 (11.6%) | 1 (5.6%) | 0 (0%) | 3 (27.3%) | 1 (33.3%) |

^a^ 9 patients with missing height or weight for body mass index (BMI) calculation were not included.

^b^ Sarcopenia was defined as skeletal muscle index (SMI) < 38.9 cm^2^/m^2^. SMI was defined as total skeletal muscle area at L3 in cm^2^ divided by height in meters squared.

Supplemental Table S7. Hazard ratios (HRs) of mortality, by body composition type

| Model | Body composition type (N = 168, 25 deaths) | | | |
| --- | --- | --- | --- | --- |
|  | Adequate | High adiposity | Low muscle | Low muscle/high adiposity |
| No. in each group | 67 | 47 | 44 | 10 |
| Deaths, No. | 8 | 7 | 8 | 2 |
| Model 1, HR (95% CI) | 1.00 (reference) | 1.19 (0.43-3.29) | 1.51 (0.57-4.04) | 1.52 (0.32-7.16) |
| Model 2, HR (95% CI) | 1.00 (reference) | 1.03 (0.37-2.90) | 2.21 (0.79-6.19) | 1.78 (0.37-8.46) |

Model 1: no covariates

Model 2: covariates include age at diagnosis, race, breast cancer stage, and tumor grade

Tertiles of TAT: 368.0 and 518.9 cm^2^; tertiles of TSM: 117.9 and 135.5 cm^2^

Supplemental Table S8. Protein marker expression level^a^ by clinical characteristic. Limited to the proteins significantly associated with body composition type shown in Table 3.

| **Tissue compartment** | Tumor | | Stroma | | | | | |
| --- | --- | --- | --- | --- | --- | --- | --- | --- |
| **Marker** | Pan-AKT | CTLA4 | INPP4B | CD8 | CD45RO | GZMB | CD20 | CD3 |
| **AJCC stage, %** |  |  |  |  |  |  |  |  |
| I | 5.65 (0.37) | 0.32 (0.57) | 2.84 (0.32) | 4.09 (0.44) | 1.84 (0.36) | 3.12 (0.38) | 0.82 (0.24) | 2.68 (0.47) |
| II | 5.64 (0.30) | 0.41 (0.46) | 2.85 (0.26) | 3.28 (0.36) | 1.56 (0.30) | 3.36 (0.31) | 0.68 (0.20) | 2.26 (0.38) |
| III | 7.16 (0.54) | 1.76 (0.82) | 3.33 (0.46) | 3.48 (0.62) | 1.37 (0.51) | 3.86 (0.54) | 1.62 (0.34) | 2.91 (0.66) |
| **Tumor grade** |  |  |  |  |  |  |  |  |
| I | 5.93 (0.73) | -0.08 (1.05) | 3.17 (0.58) | 4.15 (0.80) | 2.14 (0.64) | 4.12 (0.68) | 0.80 (0.46) | 3.04 (0.84) |
| II | 5,95 (0.31) | 0.91 (0.45) | 2.87 (0.26) | 3.57 (0.36) | 1.50 (0.29) | 3.36 (0.31) | 0.86 (0.21) | 2.53 (0.38) |
| III | 5.83 (0.37) | 0.33 (0.54) | 2.93 (0.31) | 3.44 (0.42) | 1.61 (0.34) | 3.15 (0.36) | 0.94 (0.24) | 2.34 (0.44) |
| **Tumor size** |  |  |  |  |  |  |  |  |
| <25 mm | 6.01 (0.34) | 0.43 (0.47) | 3.09 (0.27) | 3.74 (0.38) | 1.53 (0.31) | 3.34 (0.31) | 0.90 (0.21) | 2.47 (0.41) |
| ≥25 mm | 6.06 (0.39) | 1.54 (0.55) | 3.19 (0.30) | 3.81 (0.43) | 1.99 (0.35) | 3.97 (0.35) | 1.18 (0.24) | 2.89 (0.45) |
| Missing | 5.43 (0.49) | -0.51 (0.68) | 2.15 (0.39) | 2.86 (0.56) | 1.15 (0.45) | 2.38 (0.45) | 0.35 (0.31) | 1.94 (0.59) |
| **ER status** |  |  |  |  |  |  |  |  |
| Positive | 5.90 (0.24) | 0.54 (0.35) | 2.93 (0.20) | 3.68 (0.28) | 1.63 (0.22) | 3.47 (0.23) | 0.90 (0.16) | 2.60 (0.29) |
| Negative | 5.91 (0.66) | 1.01 (0.97) | 2.92 (0.53) | 2.87 (0.73) | 1.49 (0.59) | 2.63 (0.62) | 0.79 (0.42) | 1.85 (0.77) |
| **PR status** |  |  |  |  |  |  |  |  |
| Positive | 5.89 (0.25) | 0.55 (0.37) | 3.01 (0.21) | 3.64 (0.29) | 1.59 (0.23) | 3.45 (0.25) | 0.86 (0.16) | 2.51 (0.30) |
| Negative | 5.97 (0.54) | 0.80 (0.79) | 2.54 (0.43) | 3.33 (0.60) | 1.70 (0.48) | 2.99 (0.51) | 0.98 (0.34) | 2.49 (0.63) |
| **HER2 status** |  |  |  |  |  |  |  |  |
| Positive | 6.33 (0.51) | 0.88 (0.75) | 3.25 (0.43) | 3.81 (0.60) | 1.69 (0.48) | 3.80 (0.51) | 0.98 (0.34) | 3.09 (0.63) |
| Negative | 5.79 (0.25) | 0.52 (0.37) | 2.85 (0.21) | 3.53 (0.29) | 1.59 (0.23) | 3.26 (0.25) | 0.86 (0.16) | 2.38 (0.30) |

Abbreviations: AJCC, American Joint Committee on Cancer; ER; ER, estrogen receptor; PR, progesterone receptor

^a^ Least square means (standard error) estimated from mixed effect models.

Supplemental Table S9. Associations between body composition type and immune and PI3K/AKT protein expression levels in breast tumors, with additional adjustment of chemotherapy receipt

| Body composition type (compared with adequate type) | Protein marker | Tissue compartment | Estimate (log2-fold change)^a^ | P value |
| --- | --- | --- | --- | --- |
| Low muscle | Pan-AKT | Tumor | -0.79 | 0.042 |
| Low muscle | CTLA4 | Tumor | 0.54 | 0.033 |
| Low muscle | INPP4B | Stroma | 1.17 | <0.0001 |
| High adiposity | INPP4B | Stroma | 0.64 | 0.036 |
| High adiposity | CD8 | Stroma | 1.32 | 0.009 |
| High adiposity | CD45RO | Stroma | 0.98 | 0.021 |
| High adiposity | GZMB | Stroma | 0.70 | 0.08 |
| High adiposity | CD20 | Stroma | 0.59 | 0.009 |
| High adiposity | CD3 | Stroma | 1.08 | 0.056 |
| High adiposity/low muscle | Phospho-tuberin (T1462) | Tumor | -1.42 | 0.017 |
| High adiposity/low muscle | Phospho-PRAS40 (T246) | Tumor | -1.93 | 0.024 |
| High adiposity/low muscle | Phospho-PRAS40 (T246) | Stroma | -1.15 | 0.058 |
| High adiposity/low muscle | CTLA4 | Stroma | 1.25 | 0.009 |
| High adiposity/low muscle | PD-1 | Stroma | -1.43 | 0.010 |
| High adiposity/low muscle | CD14 | Stroma | -2.04 | 0.019 |
| High adiposity/low muscle | PLCG1 | Stroma | -1.21 | 0.024 |

^a^ Model adjusted for analytical batch, race, breast cancer stage, tumor grade, and chemotherapy

Supplemental Table S10. Associations of protein markers by tertiles (T) of muscle for the significant associations of the low muscle body composition type

| Protein marker (tissue compartment) | Body fatness measurements: exposure vs referent groups | Estimate (log2-fold change) | P value |
| --- | --- | --- | --- |
| Pan-AKT (tumor) | TSM T2 vs. T3 | 0.41 | 0.44 |
|  | TSM T1 vs. T3 | -0.51 | 0.48 |
|  | TSM T1 vs. T2+T3 | -0.78 | 0.057 |
|  | Low muscle vs. adequate | -0.92 | 0.043 |
| CTLA4 (tumor) | TSM T2 vs. T3 | 0.14 | 0.30 |
|  | TSM T1 vs. T3 | 0.29 | 0.33 |
|  | TSM T1 vs. T2+T3 | 0.27 | 0.30 |
|  | Low muscle vs. adequate | 0.53 | 0.059 |
| INPP4B (stroma) | TSM T2 vs. T3 | 0.09 | 0.77 |
|  | TSM T1 vs. T3 | 1.22 | 0.0009 |
|  | TSM T1 vs. T2+T3 | 1.01 | 0.0009 |
|  | Low muscle vs. adequate | 1.18 | 0.0003 |

All models adjust for analytical batch, race, breast cancer stage, and tumor grade. The total skeletal muscle (TSM) models additionally adjust for total adipose tissue tertiles.

Supplemental Table S11. Associations of protein markers for tertiles (T) of adiposity and WC for the significant associations of the high adiposity body composition type

| Protein marker (tissue compartment) | Body fatness measurements: exposure vs referent groups | Estimate (log2-fold change) | P value |
| --- | --- | --- | --- |
| INPP4B (stroma) | TAT T2 vs. T1 | 0.42 | 0.13 |
|  | TAT T3 vs. T1 | 0.86 | 0.017 |
|  | TAT T3 vs. T2+T1 | 0.43 | 0.16 |
|  | High adiposity vs. adequate | 0.64 | 0.058 |
|  | WC T2 vs. T1 | 0.03 | 0.94 |
|  | WC T3 vs. T1 | 0.48 | 0.18 |
| CD8 (stroma) | TAT T2 vs. T1 | 0.16 | 0.76 |
|  | TAT T3 vs. T1 | 0.37 | 0.56 |
|  | TAT T3 vs. T2+T1 | 0.70 | 0.18 |
|  | High adiposity vs. adequate | 1.16 | 0.043 |
|  | WC T2 vs. T1 | 0.06 | 0.92 |
|  | WC T3 vs. T1 | 0.27 | 0.62 |
| CD45RO (stroma) | TAT T2 vs T1 | 0.53 | 0.21 |
|  | TAT T3 vs. T1 | 0.94 | 0.07 |
|  | TAT T3 vs. T2+T1 | 0.76 | 0.07 |
|  | High adiposity vs. adequate | 0.95 | 0.043 |
|  | WC T2 vs. T1 | 0.03 | 0.96 |
|  | WC T3 vs. T1 | 0.30 | 0.65 |
| GZMB (stroma) | TAT T2 vs. T1 | 0.10 | 0.81 |
|  | TAT T3 vs. T1 | 0.28 | 0.58 |
|  | TAT T3 vs. T2+T1 | 0.15 | 0.72 |
|  | High adiposity vs. adequate | 0.63 | 0.17 |
|  | WC T2 vs. T1 | 0.34 | 0.48 |
|  | WC T3 vs. T1 | 0.41 | 0.36 |
| CD20 (stroma) | TAT T2 vs. T1 | 0.004 | 0.99 |
|  | TAT T3 vs. T1 | 0.20 | 0.54 |
|  | TAT T3 vs. T2+T1 | 0.33 | 0.20 |
|  | High adiposity vs. adequate | 0.61 | 0.031 |
|  | WC T2 vs. T1 | 0.23 | 0.42 |
|  | WC T3 vs. T1 | 0.45 | 0.09 |
| CD3 (stroma) | TAT T2 vs. T1 | 0.21 | 0.72 |
|  | TAT T3 vs. T1 | 0.35 | 0.62 |
|  | TAT T3 vs. T2+T1 | 0.60 | 0.29 |
|  | High adiposity vs. adequate | 1.07 | 0.09 |
|  | WC T2 vs. T1 | 0.06 | 0.93 |
|  | WC T3 vs. T1 | 0.10 | 0.86 |

All models adjust for analytical batch, race, breast cancer stage, and tumor grade. The total adipose tissue (TAT) and waist circumference (WC) models additionally adjust for total skeletal muscle tertiles.

Supplemental Table S12. Associations of protein markers for body mass index and high adiposity for the markers significantly associated with the high adiposity body composition type, restricted to participants with data on body mass index (N= 40)

| Protein marker (tissue compartment) | Body fatness measurement: exposure vs. referent groups | Estimate (log2-fold change) | P value |
| --- | --- | --- | --- |
| INPP4B (stroma) | High adiposity vs. adequate | 0.63 | 0.07 |
|  | Overweight vs. normal weight | -0.40 | 0.36 |
|  | Obesity vs. normal weight | -0.15 | 0.76 |
| CD8 (stroma) | High adiposity vs. adequate | 1.97 | 0.001 |
|  | Overweight vs. normal weight | -0.47 | 0.48 |
|  | Obesity vs. normal weight | 0.29 | 0.71 |
| CD45RO (stroma) | High adiposity vs. adequate | 1.51 | 0.0025 |
|  | Overweight vs. normal weight | 0.13 | 0.81 |
|  | Obesity vs. normal weight | 0.16 | 0.80 |
| GZMB (stroma) | High adiposity vs. adequate | 0.47 | 0.28 |
|  | Overweight vs. normal weight | -0.76 | 0.12 |
|  | Obesity vs. normal weight | -0.39 | 0.48 |
| CD20 (stroma) | High adiposity vs. adequate | 0.64 | 0.0057 |
|  | Overweight vs. normal weight | -0.49 | 0.05 |
|  | Obesity vs. normal weight | 0.06 | 0.83 |
| CD3 (stroma) | High adiposity vs. adequate | 1.68 | 0.0124 |
|  | Overweight vs. normal weight | -0.28 | 0.70 |
|  | Obesity vs. normal weight | 0.48 | 0.57 |

All models adjust for analytical batch, race, breast cancer stage, and tumor grade.

Supplemental Table S13. Statistical power of protein markers in the tumor and stromal compartments

|  | Marker | Power | |
| --- | --- | --- | --- |
|  |  | Tumor | Stromal |
| 1 | CD8 | 0.170 | 0.545 |
| 2 | GZMB | 0.125 | 0.625 |
| 3 | CD11c | 0.120 | 0.610 |
| 4 | PD-L1 | 0.330 | 0.435 |
| 5 | CTLA4 | 0.645 | 0.615 |
| 6 | Beta-2-microglobulin | 0.340 | 0.420 |
| 7 | CD20 | 0.120 | 0.640 |
| 8 | CD68 | 0.140 | 0.420 |
| 9 | SMA | 0.100 | 0.490 |
| 10 | Fibronectin | 0.205 | 0.385 |
| 11 | CD45 | 0.105 | 0.540 |
| 12 | PanCk | 0.155 | 0.165 |
| 13 | HLA-DR | 0.370 | 0.525 |
| 14 | Ki-67 | 0.375 | 0.485 |
| 15 | PD-1 | 0.360 | 0.500 |
| 16 | CD56 | 0.230 | 0.325 |
| 17 | CD3 | 0.170 | 0.475 |
| 18 | CD4 | 0.110 | 0.460 |
| 19 | CD45RO | 0.315 | 0.450 |
| 20 | FOXP3 | 0.265 | 0.200 |
| 21 | CD163 | 0.395 | 0.530 |
| 22 | CD66b | 0.260 | 0.235 |
| 23 | CD14 | 0.240 | 0.695 |
| 24 | FAP-alpha | 0.400 | 0.360 |
| 25 | CD34 | 0.215 | 0.670 |
| 26 | Pan-AKT | 0.500 | 0.385 |
| 27 | Phospho-GSK3B (S9) | 0.365 | 0.380 |
| 28 | PLCG1 | 0.350 | 0.590 |
| 29 | Phospho-Tuberin (T1462) | 0.700 | 0.315 |
| 30 | Phospho-AKT1 (S473) | 0.435 | 0.465 |
| 31 | Phospho-GSK3A (S21)/Phospho-GSK3B (S9) | 0.360 | 0.325 |
| 32 | MET | 0.285 | 0.510 |
| 33 | Phospho-PRAS40 (T246) | 0.565 | 0.560 |
| 34 | INPP4B | 0.495 | 0.960 |
| 35 | PTEN | 0.265 | 0.740 |
| 36 | PR | 0.305 | 0.510 |
| 37 | ER-alpha | 0.405 | 0.485 |
| 38 | EpCAM | 0.290 | 0.210 |
| 39 | Her2 | 0.595 | 0.485 |
| 40 | MART1 | 0.190 | 0.275 |
| 41 | Bcl-2 | 0.470 | 0.525 |
| 42 | S100B | 0.165 | 0.525 |
| 43 | NY-ESO-1 | 0.405 | 0.490 |

Supplemental Table S14. Characteristics of patients with tumor samples on tissue microarrays (TMAs) vs. patients without or with ineligible tumor samples vs. patient records from the Tumor Registry at UF Health

| **Characteristic** | **Patients with tumor samples on TMA^a^** | **Patients without or with ineligible tumor samples** | **Tumor Registry at UF Health^b^** |
| --- | --- | --- | --- |
|  | **Mean (SD) or No. (%)** | **Mean (SD) or No. (%)** | **Mean (SD) or No. (%)** |
| **N** | 57 | 527 | 2223 |
| Age at diagnosis (years) | 57.1 (10.7) | 58.3 (11.2) | 58.6 (12.6) |
| **Race** |  |  |  |
| White | 48 (84.2) | 412 (79.4) | 1809 (82.3) |
| Black | 9 (15.8) | 107 (20.6) | 313 (14.3) |
| Asian | 0 | 0 | 35 (1.6) |
| PI/AI | 0 | 0 | 2 (0.1) |
| Others | 0 | 0 | 38 (1.7) |
| Missing | 0 | 8 | 26 |
| **AJCC clinical stage** |  |  |  |
| 0 | 0 | 5 (1.2) | 430 (21.7) |
| I | 21 (36.8) | 167 (41.5) | 901 (45.6) |
| II | 27 (47.3) | 127 (31.6) | 401 (20.3) |
| III | 9 (15.8) | 59 (14.7) | 105 (5.3) |
| IV | 0 (0) | 44 (10.9) | 141 (7.1) |
| Missing | 0 | 125 | 245 |
| **Tumor grade** |  |  | NA |
| I | 5 (8.8) | 50 (13.5) |  |
| II | 29 (50.9) | 145 (39.1) |  |
| III | 23 (40.3) | 176 (47.4) |  |
| Missing | 0 | 156 |  |
| **Tumor size** |  |  | NA |
| <25 mm | 26 (56.5) | 165 (58.1) |  |
| ≥25 mm | 10 (43.5) | 119 (41.9) |  |
| Missing | 11 | 243 |  |
| **ER status** |  |  |  |
| Positive | 49 (86.0) | 299 (76.3) | 1724 (82.8) |
| Negative | 8 (14.0) | 93 (23.7) | 357 (17.2) |
| Missing | 0 | 135 | 142 |
| **PR status** |  |  |  |
| Positive | 46 (80.7) | 264 (68.2) | 1538 (74.5) |
| Negative | 11 (19.3) | 123 (31.8) | 526 (25.5) |
| Missing | 0 | 140 | 159 |
| **HER2 status** |  |  |  |
| Positive/equivocal | 10 (17.9) | 68 (20.1) | 284 (19.7) |
| Negative | 46 (82.1) | 270 (79.9) | 1158 (80.3) |
| Missing/test not done | 1 | 189 | 781 |
| **Year of CT scan** |  |  | NA |
| 2011 | 0 | 16 (3.4) |  |
| 2012 | 1 (1.9) | 59 (12.4) |  |
| 2013 | 13 (24.1) | 83 (17.5) |  |
| 2014 | 8 (14.8) | 59 (12.4) |  |
| 2015 | 6 (11.1) | 67 (14.1) |  |
| 2016 | 3 (5.6) | 81 (17.1) |  |
| 2017 | 9 (16.7) | 61 (12.8) |  |
| 2018 | 12 (22.2) | 27 (5.7) |  |
| 2019 | 2 (3.7) | 20 (4.2) |  |
| 2020 | 0 | 2 (0.4) |  |
| Missing | 3 | 52 |  |
| **Year of diagnosis** |  |  |  |
| Before 2011 | 0 | 98 (20.9) | 519 (23.4) |
| 2011 | 3 (5.3) | 35 (7.5) | 210 (9.5) |
| 2012 | 3 (5.3) | 41 (8.8) | 210 (9.5) |
| 2013 | 12 (21.1) | 41 (8.8) | 235 (10.6) |
| 2014 | 4 (7.0) | 53 (11.3) | 240 (10.8) |
| 2015 | 7 (12.3) | 50 (10.7) | 263 (11.8) |
| 2016 | 4 (7.0) | 53 (11.3) | 257 (11.6) |
| 2017 | 8 (14.0) | 39 (8.3) | 277 (12.5) |
| 2018 | 10 (17.5) | 32 (6.8) | 12 (0.5) |
| 2019 | 6 (10.5) | 15 (3.2) | NA |
| After 2019 | 0 | 11 (2.4) | NA |
| Missing | 0 | 59 | 0 |

Abbreviations: AJCC, American Joint Committee on Cancer; CT, computed tomography; ER; ER, estrogen receptor; NA, not available; PI/AI, Pacific Islanders/American Indians; PR, progesterone receptor

^a^ Including 52 patients who had GeoMx Digital Spatial Profiler complete data and 5 patients who had tumor tissue on tissue microarrays but the tissue failed during the assay.

^b^ Restricted to females and analytic cases (class of case 00-22, patients who were diagnosed or administered any of their first course of treatment at the accessioning facility after the registry’s reference date are analytic), 2008 to January 2018.

Supplemental Figure S1. Flowchart of study participants and tumor tissue sample selection

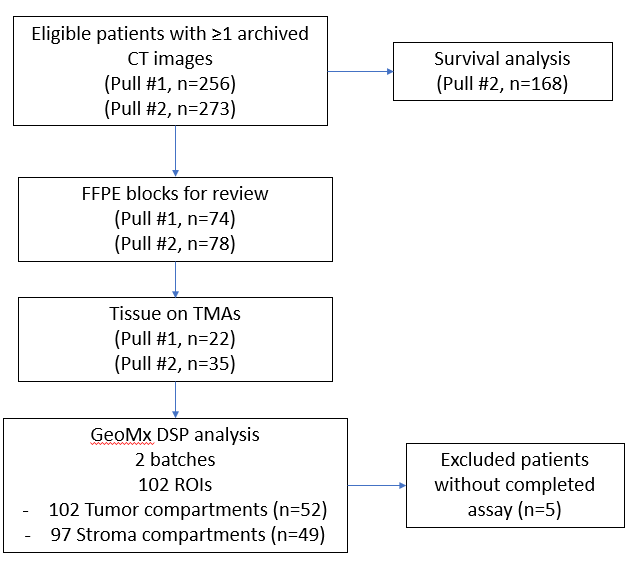

Abbreviations: CT, computed tomography; DSP, Digital Spatial Profiler; FFPE, formalin-fixed, paraffin-embedded; ROIs, regions of interest; TMA, tissue microarray.

Supplemental Figure S2. Total adipose and muscle tissue areas by body composition phenotype.

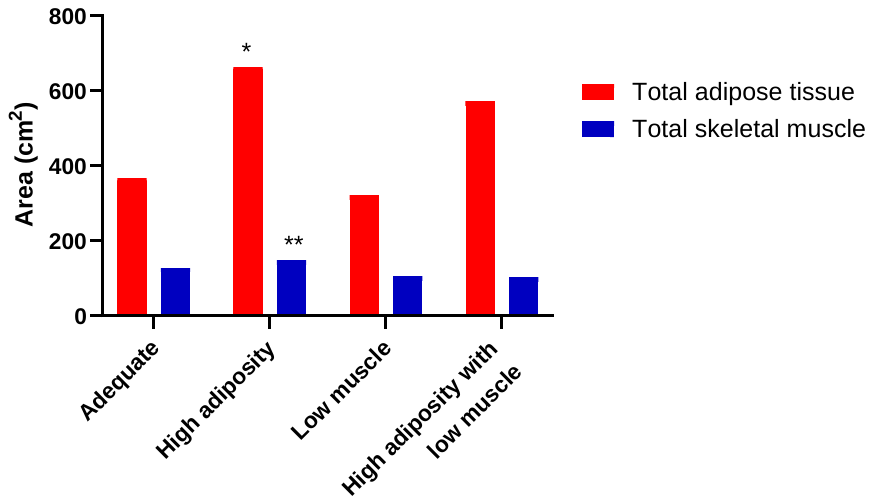

* P<0.05 (t-tests) compared with the adequate and the low muscle groups.

** P<0.05 (t-tests) compared with the adequate, the low muscle, and the high adiposity with low muscle groups

Supplemental Figure S3. Percentages of different densities of muscle in total skeletal muscle area by body composition phenotype. Abbreviations: VHDM, very high-density muscle; HDM, high-density muscle; NDM, normal-density muscle; LDM, low-density muscle; VLDM, very low-density muscle

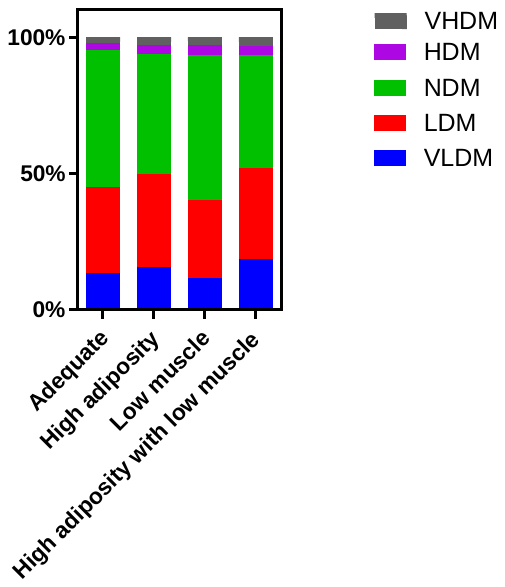

P>0.05 for each muscle type between body composition groups (two proportion Z-tests)

Supplemental Figure S4. Box and whisker plots showing levels of estrogen receptor (ER)-alpha, progesterone receptor (PR), and HER2 proteins in the epithelium compartment measured using a GeoMx Digital Spatial Profiler plotted by ER, PR, and HER2 status obtained from pathology reports (immunohistochemistry). Panel A: ER; Panel B: PR; Panel C: HER2. Horizontal lines indicate medians; outer edges of boxes, Q1 and Q3 values; error bars, minimum and maximum values. Between-group differences were examined using linear mixed models (P=0.20 for ER, P=0.13 for PR, and P<0.001 for HER2).

| A | 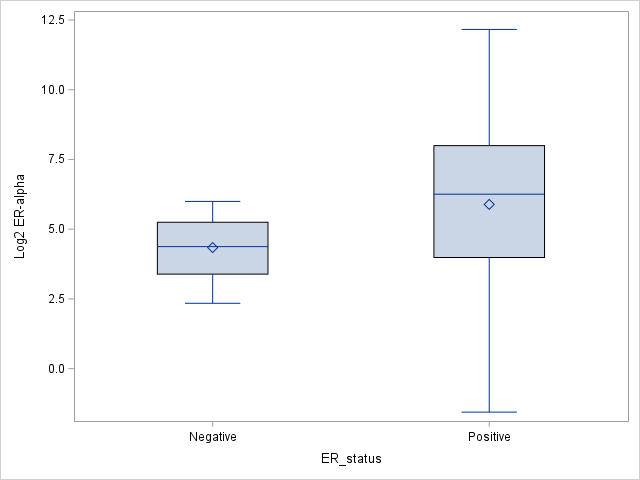 |
| --- | --- |
| B | 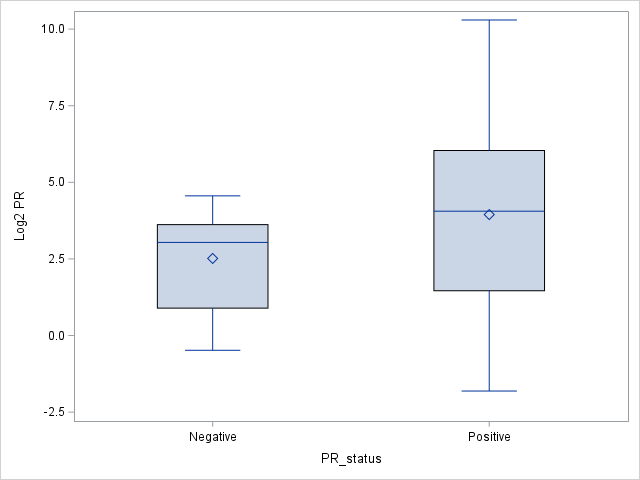 |
| C | 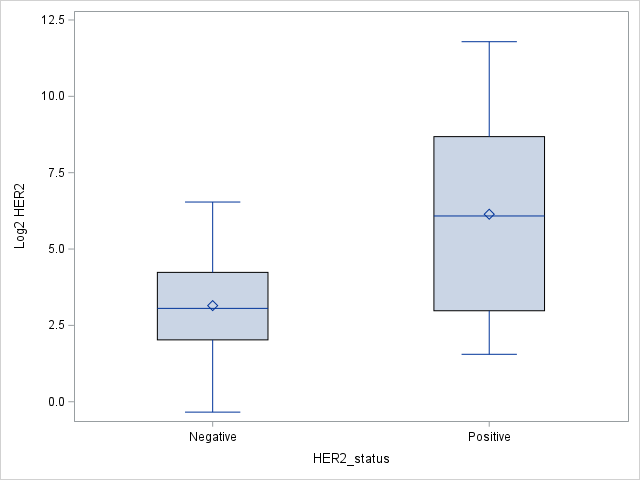 |

Supplemental Figure S5. Heatmap by body composition type for proteins in tumor (A) epithelium and (B) stroma

| A | 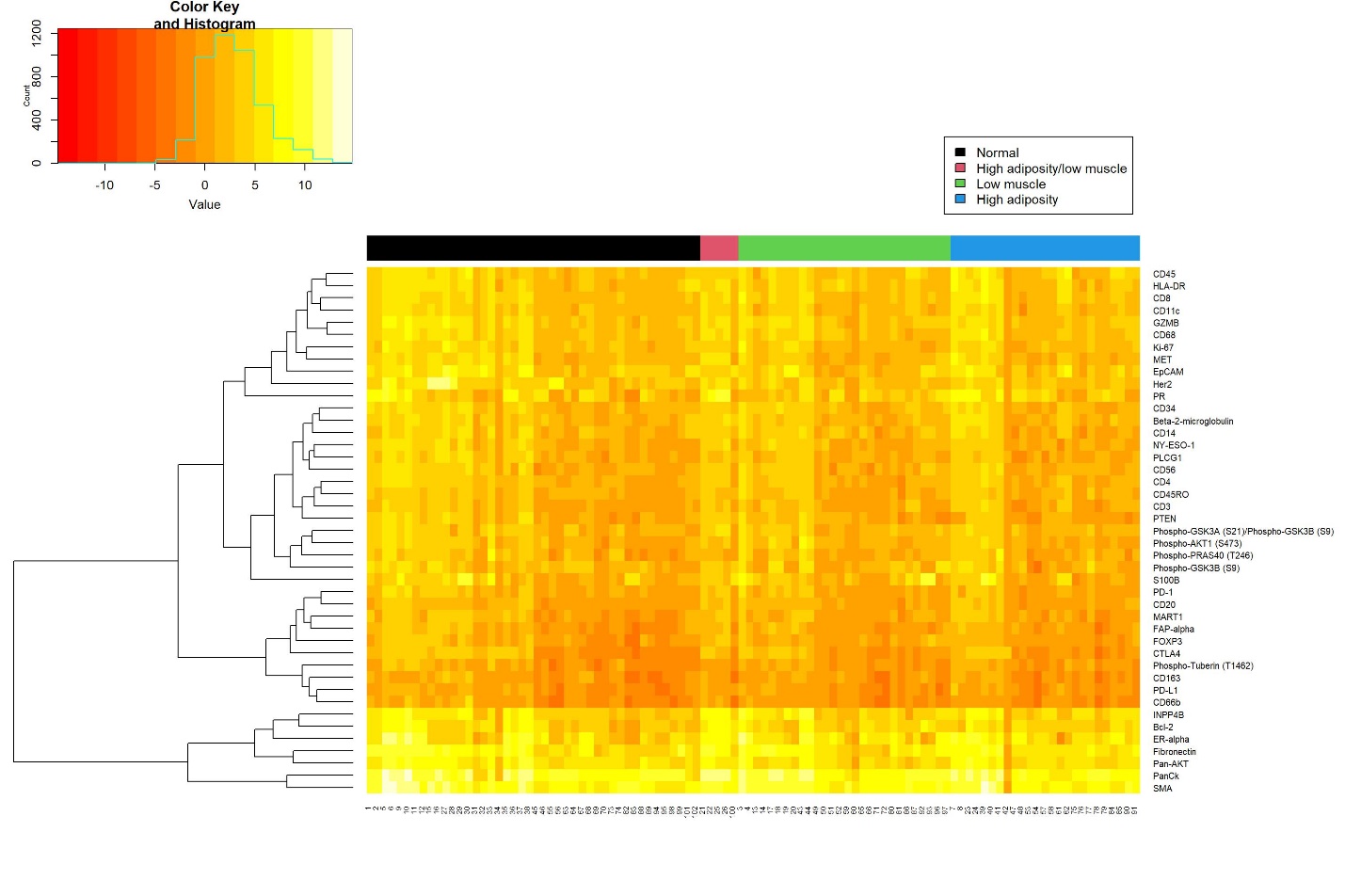 |
| --- | --- |
| B | 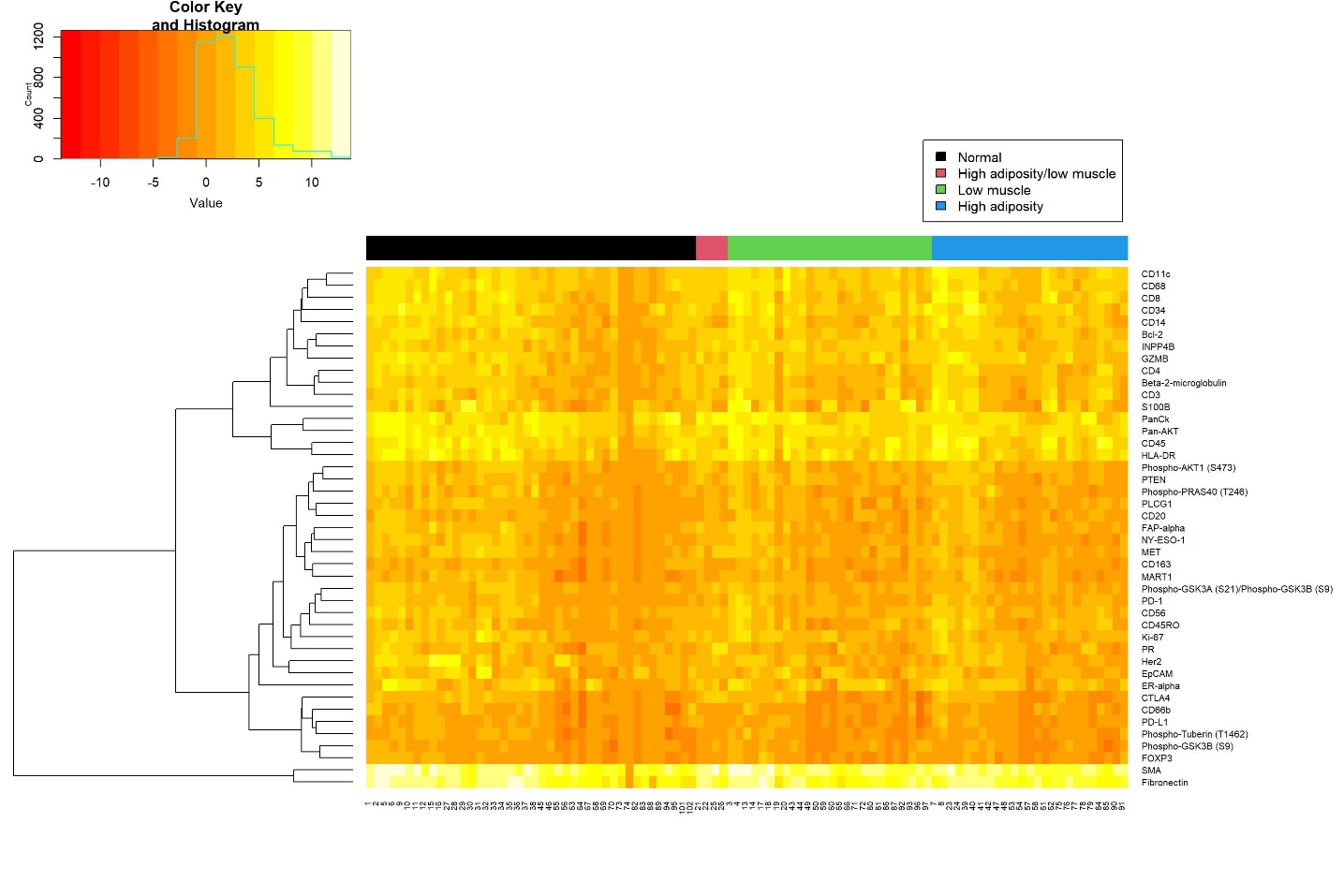 |

Supplemental Figure S6. Survival curves for (A) phospho-tuberin T1462 in luminal A breast cancer, (B) phospho-tuberin T1462 in luminal B breast cancer, (C) phospho-PRAS40 T246 in Luminal A breast cancer, and (D) phospho-PRAS40 T246 in Luminal B breast cancer. End points are breast cancer-specific survival. High and low levels of the proteins are derived by the optimal cutoff method. Source: The Cancer Protein Atlas: [TRGAted (shinyapps.io)](https://nborcherding.shinyapps.io/TRGAted/).

| A | 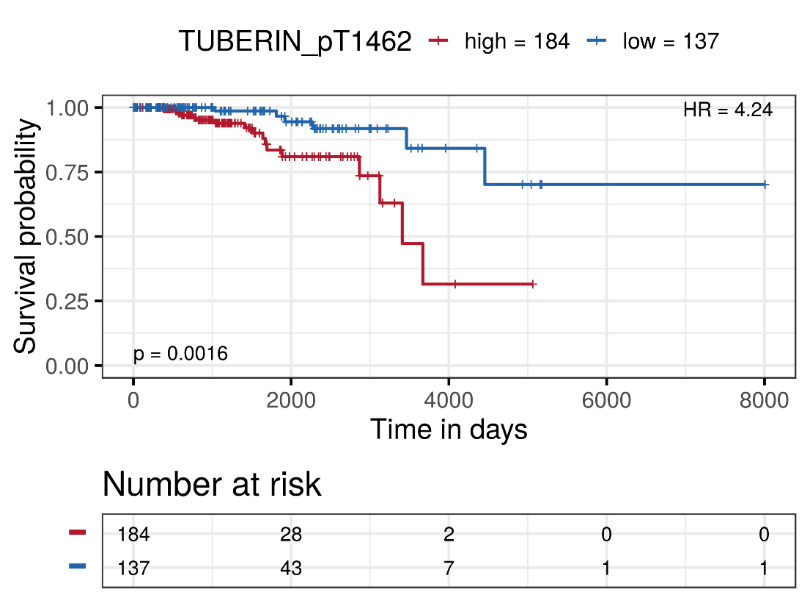 |
| --- | --- |
| B | 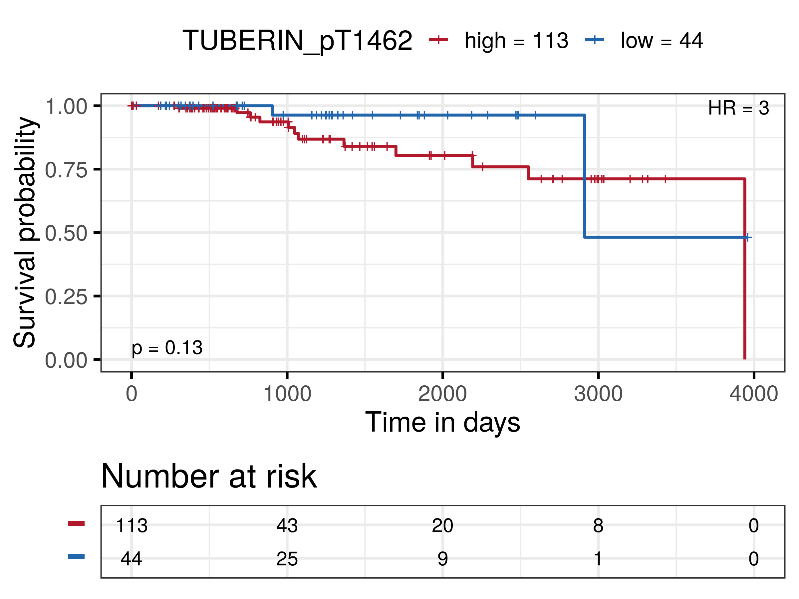 |
| C | 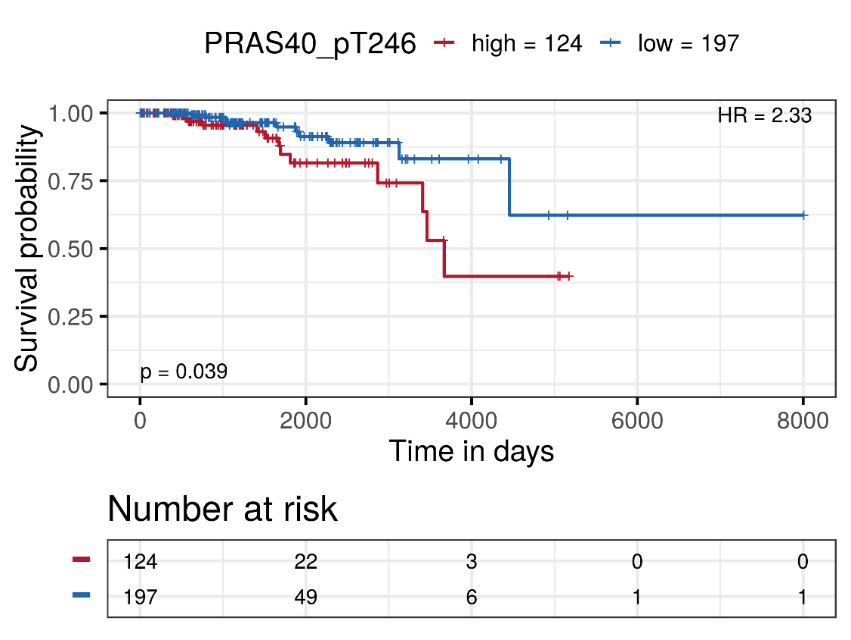 |
| D | 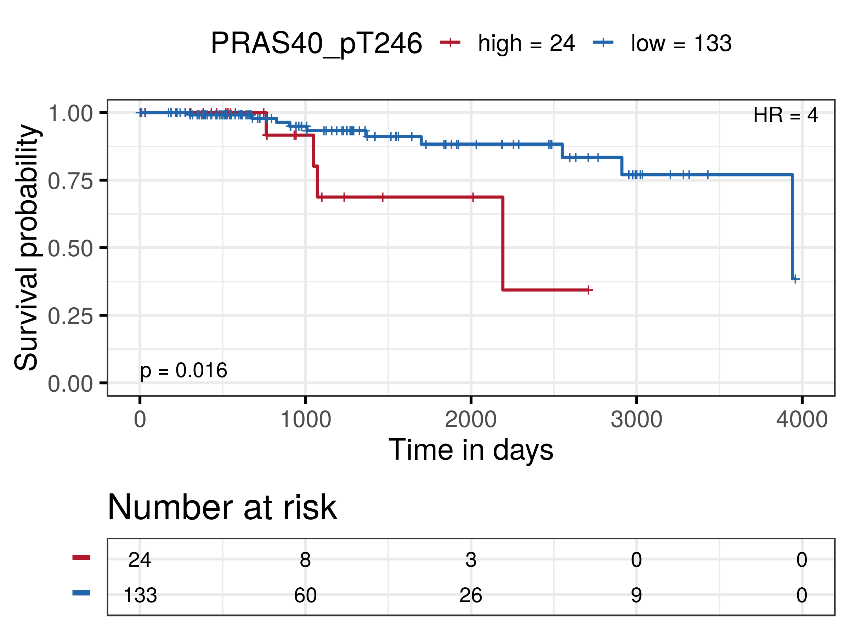 |
